# Analysis and Applications of Adaptive Group Testing Methods for COVID-19

**DOI:** 10.1101/2020.04.05.20050245

**Authors:** Cassidy Mentus, Martin Romeo, Christian DiPaola

**Affiliations:** University of California, Los Angeles; Hollings Cancer Center, Medical University of South Carolina; UMass Memorial

## Abstract

Testing strategies for Covid-19 to maximize number of people tested are urgently needed. Recently, it has been demonstrated that RT-PCR has the sensitivity to detect one positive case in a mixed sample of 32 cases [12], In this paper we propose adaptive group testing strategies based on generalized binary splitting (GBS) [5], where we restrict the group test to the largest group that can be used. The method starts by choosing a group from the population to be tested, performing a test on the combined sample from the entire group, and progressively splitting the group further into subgroups. Compared to individual testing at 4% prevalence, we save 74%; at 1% we save 91%; and at .1% we save 98% of tests. We analyze the number of times each sample is used and show that the method is still efficient if we resort to testing a case individually if the sample is running low.

In addition we recommend clinical screening to filter out individuals with symptoms and show this leaves us with a population with lower prevalence. Our approach is particularly applicable to vulnerable confined populations such as nursing homes, prisons, military ships and cruise ships.

## 1 Introduction

Testing capacity for COVID-19 is still too scarce to meet the needs to meet global health needs. Confined populations are at particular risk for rapid contagion. They may include those who reside in facilities such as prisons, ships, military units and nursing homes. The goal of this paper is to increase the capacity to identify asymptomatic carriers of COVID-19 by applying group testing methods. Current practice typically involves a “one-patient-one-test” strategy. Recently the potential to detect COVID-19 RNA in a mixture of samples from individuals has been validated [12] using RT-PCR. Group testing was first studied mathematically in 1943 during World War II to test large groups of US servicemen for syphillis prior to deployment [3]. Now, just as in the WWII era, large scale testing is necessary.

COVID-19 is a highly contagious disease that can lead to pneumonia, acute respiratory distress syndrome (ARDS) and death. Clinical symptoms and phyiscal exam features may include fever, cough, shortness of breath, malaise, lethargy, ageusia, anosmia and gastrointestinal (GI) symptoms. Besides this disease’s potential lethality, it is highly contagious.

Recently the potential of group testing using RT-PCR for finding SARS-CoV-2 RNA has been demonstrated [12]. In Europe, group-testing is currently being considered as a possible important step to totally cut-off the possibility of a resurgence as our other methods succeed in driving the rate of infection down [4]. In Luxembourg, massive testing to find asymptomatic carriers is underway and group testing can fill in the gaps in testing asymptomatic positive cases. In Hungary, there are plans to potentially test every person in the entire country using group-tests [6].

There is renewed interest in the practical applications of group testing algorithms. In 18 a non-adaptive group testing method and practical applications are explored. We describe adaptive group testing methods based on generalized binary splitting (GBS)[5]. We provide an algorithmic specification for subdividing groups and account for the limited test accuracy and the possibility that the an individual’s sample size constrains the number of tests. We explain how prescreening symptomatic cases and separately testing them can dramatically improve performance. Pre-screening reduces the prevalence in the test groups. This approach is relevant for large scale populations and particularly for confined groups.

## 2 Application of group tests to confined populations

This application utilizes a primary clinical screening step to ensure that the tested population is composed of asymptomatic COVID-19 (−) and (+) patients. This ensures the lowest prevalence of disease in the population and enhances the efficacy of the method.

Clinical screening should first take place in the selected population to screen out as many potential positive patients as possible before administering the test. All patients with a history of cough, shortness of breath, nausea, gastrointestinal symptoms, fever, malaise, lethargy, recent contact with positive COVID-19 patients, or having a physical exam consistent with those findings should be segregated out of the population.

Clinical screening will decrease the prevalence of COVID-19 carriers in the test population if the probability of being asymptomatic given COVID-19 (+) is less than the proportion of the population that is asymptomatic (regardless of COVID-19 (+) or (−)). We demonstrate this by proving the following claim.

*Claim*. The sub-population not showing any symptoms will have a lower proportion of asymptomatic carriers when the proportion of people not showing any symptoms in our group is less than the estimated proportion of COVID-19 carriers who are asymptomatic.

We can prove this through a couple of applications of Bayes’ theorem. We refer to the event that an individual does not show symptoms as “no symptoms”. The event that an individual has symptoms that are signs of COVID-19 is referred to as “symptoms” regardless of whether they carry the disease. We denote the event of carrying the COVID-19 virus as “COVID-19”. Then an asymptomatic carrier is referred to as “no symptoms and COVID-19”. Let *P*(*A*) denote the probability of an event *A* occuring. Written in terms of probabilities, screening will help when

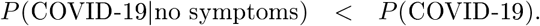

Re-writing the left side of the inequality, we get

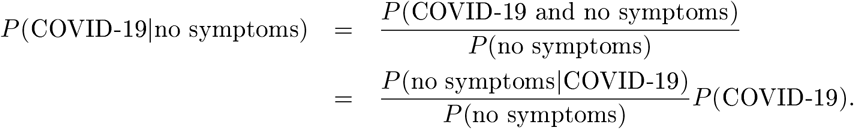

Therefore it is necessary and sufficient for 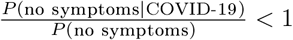, which is the same as *P* (no symptoms|COVID-19) < *P* (no symptoms). This is the probability of being asymptomatic given that they have COVID-19 for the population we are testing.

## 3 Overview of group test methods

If a population has a low prevalence of COVID-19 then it is likely for groups of individuals to not have any positive cases. Therefore it is often the case that one test of the mixture of their samples is all that is needed to determine that they are all negative.

Otherwise, a positive result on the combined samples indicates that there are some positive cases. We are therefore able to design a strategy to use less tests to determine whether each individual is positive or negative for the disease by testing mixtures of samples from groups.

To sufficiently identify negative cases and positve cases the groups must be large enough to balance finding many negative cases with one test, and locating some positive cases. Step one of the group test method is:

**1**. A group size is chosen so that the frequency that a group has only (−) cases (figure 3.1) is roughly equal to the probability we find at least one (+) case in the group.

**Figure 1.1:**
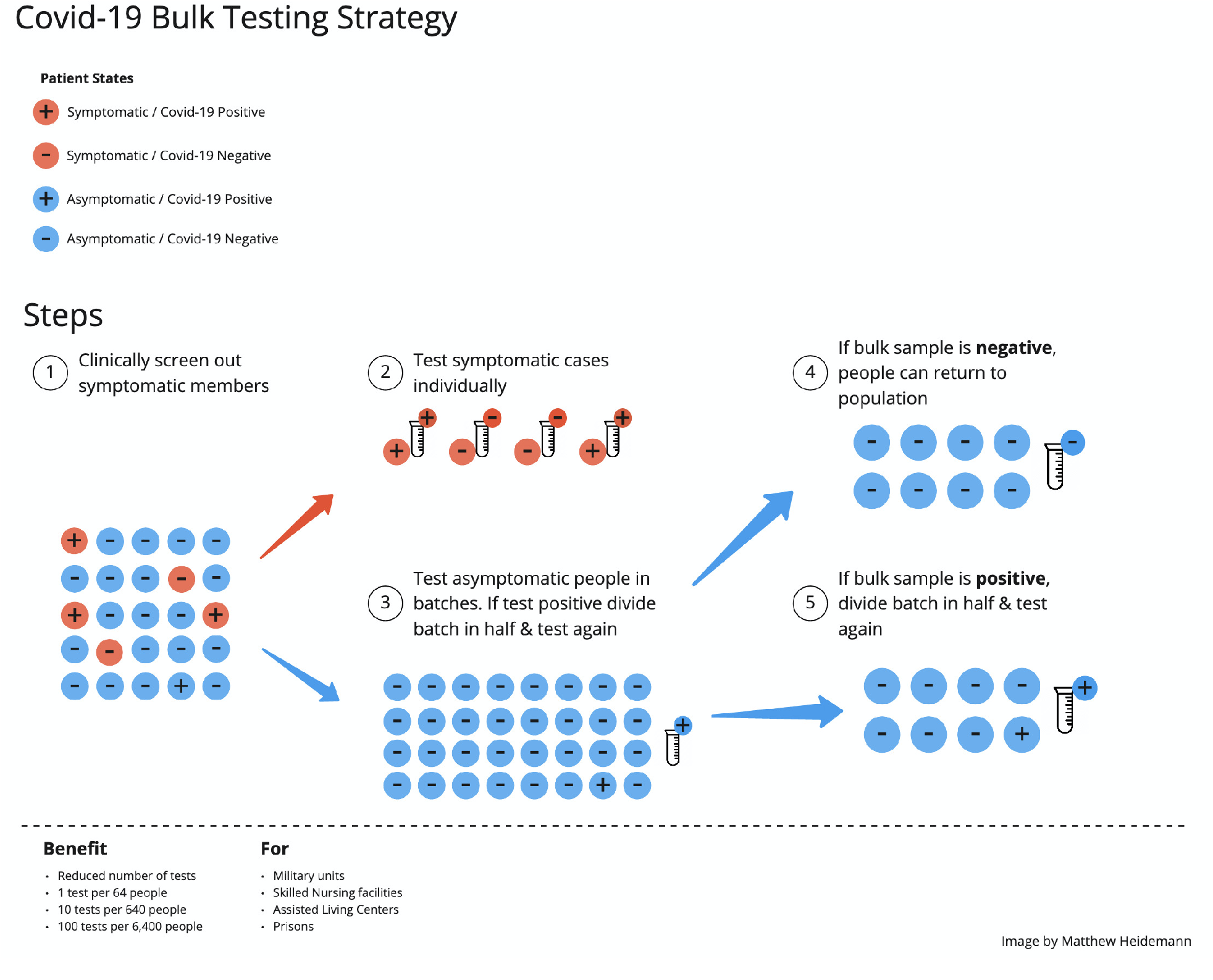
(Image credit: 1atthew Heidemann) Our group testing method consists of first clinically screening out symptomatic individuals. This will lower the prevalence in the test population. Our group testing is especially effective when we can assume the prevalence is uniform over the whole population. Therefore, it is especially applicable to confined or cohesive populations. We split the asymptomatic individuals up into groups for which we mix the samples from each individual and test them. A negative result will confirm many negative cases. We subsequently divide groups and test the mixtures of samples for positive results. This, we show, conserves the number of tests and, as a result, the time spent testing. Using numerical experiments, we also show that our methods can be performed without running out of samples from passing through too many rounds.

**Figure 2.1:**
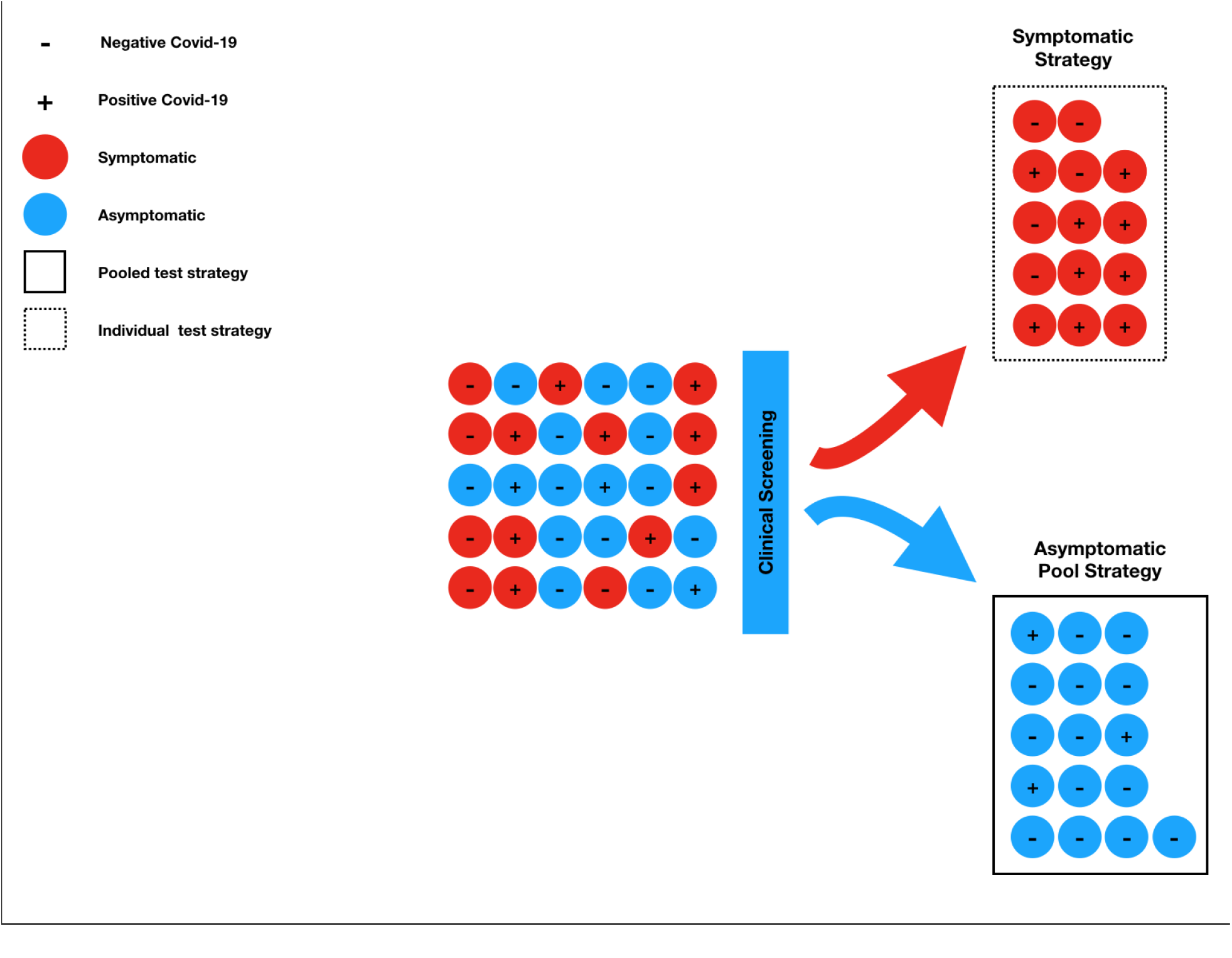
Diagram depicting the application of screening.

**Figure 3.1:**
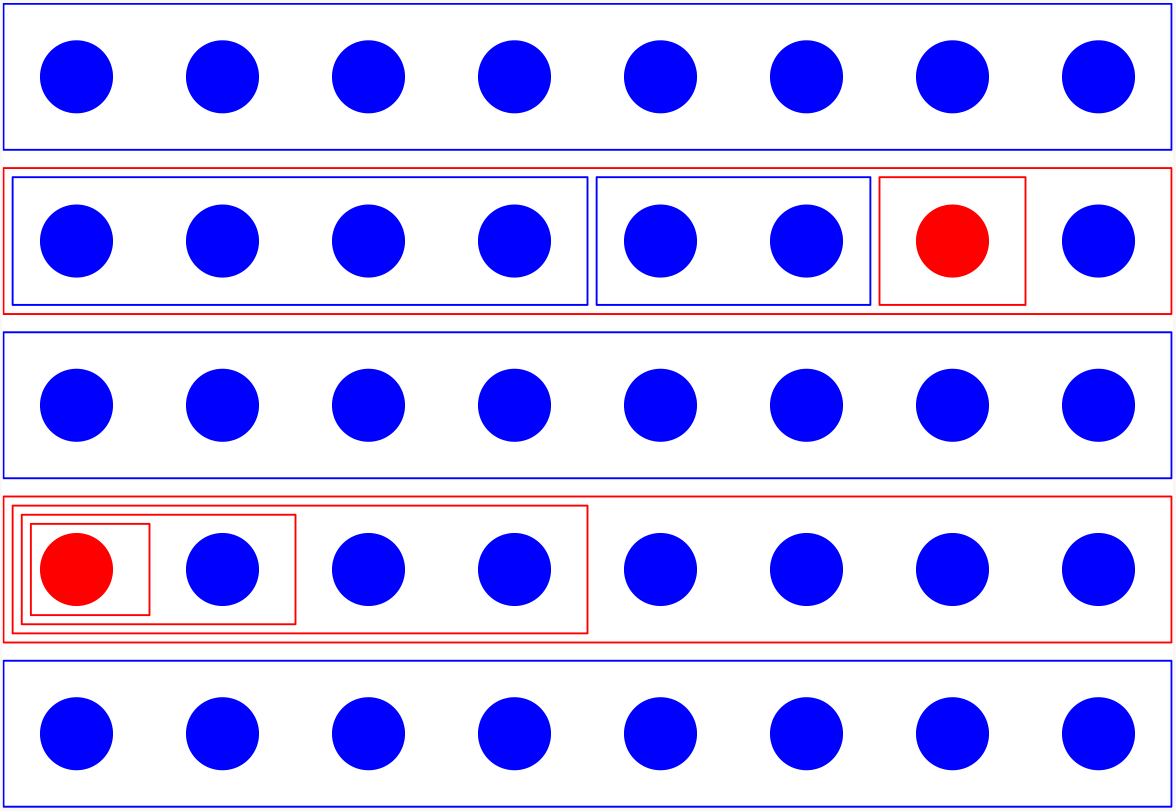
An example of a population with low prevalence. For this example only, a red circle represents a case (+) for COVID-19 and a blue circle represents a case (−) for COVID-19. The rectangles represent testing a mixture of samples from the individuals in the group. The maximum group size in this example is 8. A blue frame represents testing (−) for COVID-19 and a red frame represents testing (+). Two of the group tests of 8 are positive. When this happens, a type of binary search is used to find one positive case. In this example 32 cases are confirmed either (+) or (−) using 11 tests. Some cases are left un-determined for future rounds of testing.

**Figure 3.2:**
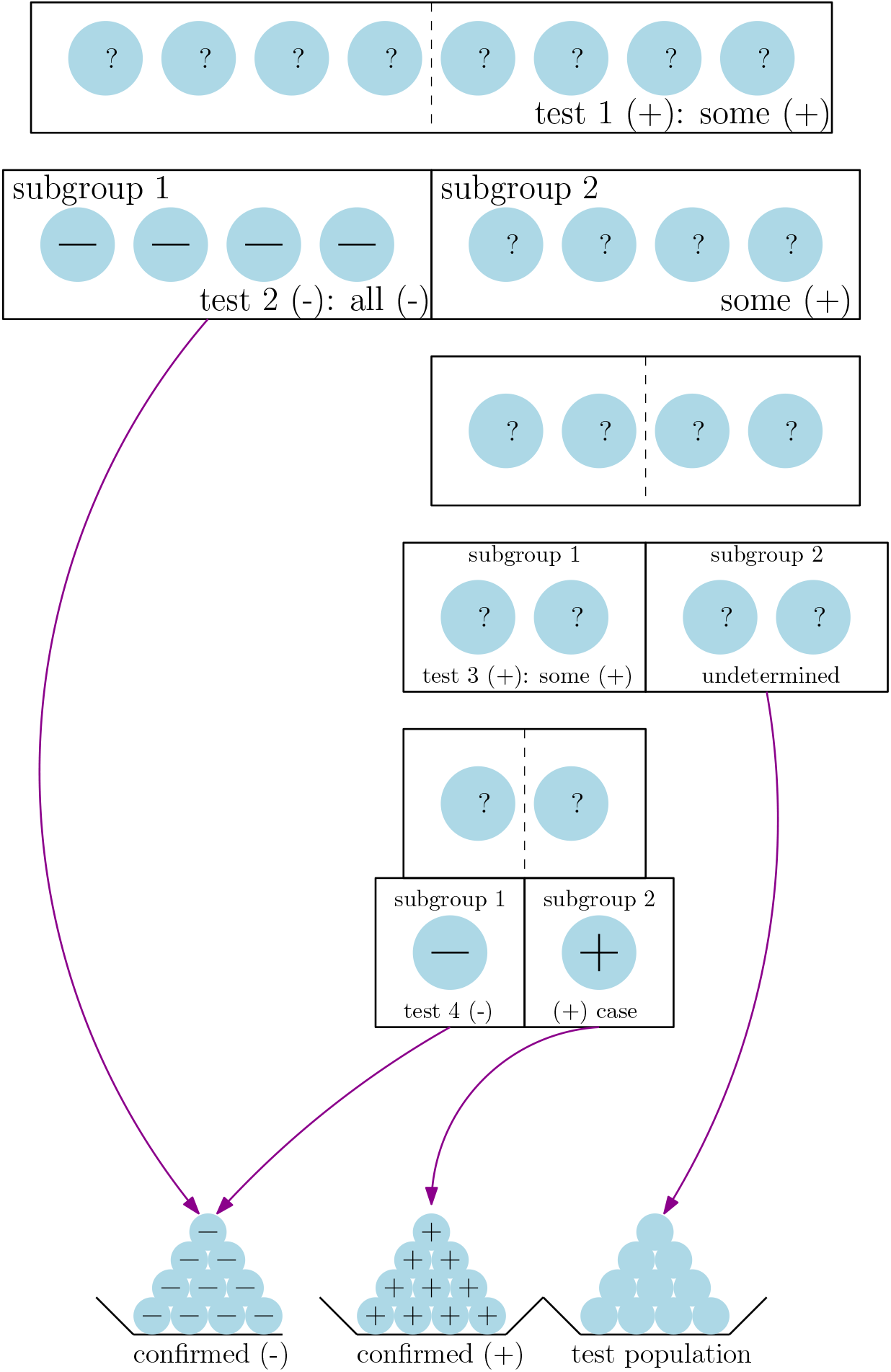
The result of testing the combined samples in the group is positive. Therefore split the group into two subgroups of equal size. Test subgroup 1. The result is (−). Therefore every case in subgroup 1 is negative. The (+) cases have to be in subgroup 2. We divide subgroup 2 into two subgroups and pass down the labels subgroup 1 and 2. The combined samples from subgroup 1 test positive. Subgroup 2 will not be tested in this round of binary search, so we return the cases to the test population. Finally split the remaining subgroup into two individual cases. Testing the first case reveals that they are (−). The second case is then (+). In total: 5 (−) cases and 1 (+) case are revealed using 4 tests.

**Figure 9.1:**
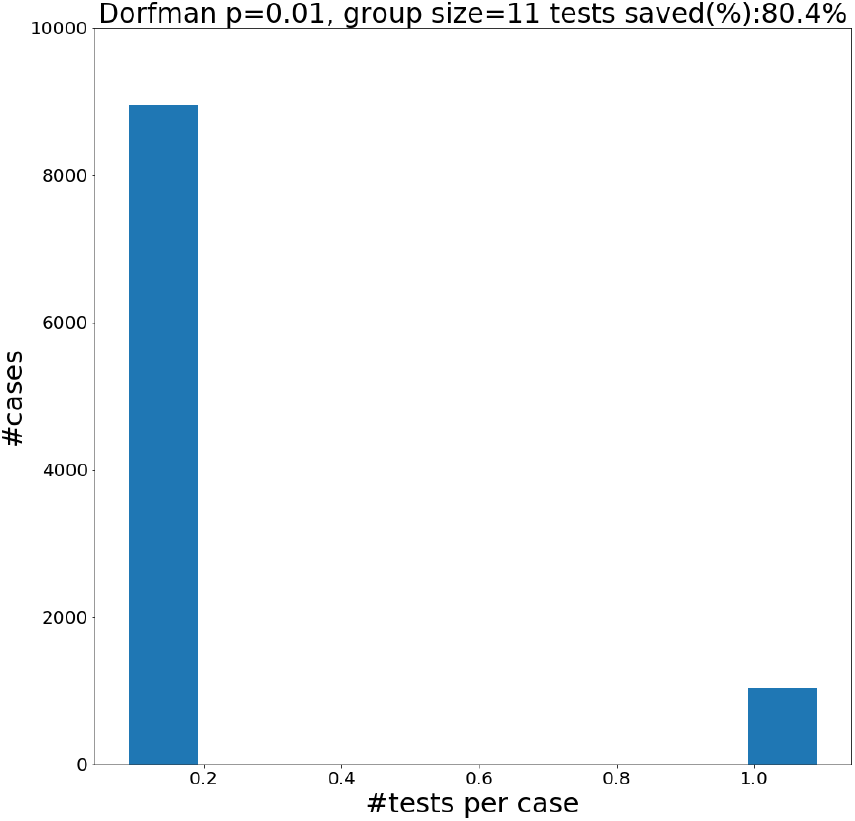
Dorfman’s method with group size *b* takes either 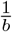 tests per case to confirm a negative case when it is in a group of comprised of *b* negatives, or 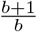 tests per case if the result is positve. If there are some positive cases in the group, in addition to the test on the mixture of samples from the whole group, a test is applied to each individual in the group. This leads the average number of tests per individual to be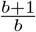.

**Table 3.1:** Group size specified for the divide and test method. By DT*g* we mean to apply divide and test, using the minimum of *g* and the group size for divide and test (e.g. DT32 will use a group of size 32 if DT uses a group of size 64). We study limited group sizes to explore how many tests can be saved depending on the limitations of the instruments.

| | $p = .001$ | .01 | .02 | .03 | .04 | .05 | .1 | .15 | .2 | .25 |
| --- | --- | --- | --- | --- | --- | --- | --- | --- | --- | --- |
| DT | 512 | 64 | 32 | 32 | 16 | 16 | 8 | 4 | 4 | 2 |
| DT4 | 4 | 4 | 4 | 4 | 4 | 4 | 4 | 4 | 4 | 2 |
| DT8 | 8 | 8 | 8 | 8 | 8 | 8 | 8 | 4 | 4 | 2 |
| DT16 | 16 | 16 | 16 | 16 | 16 | 16 | 8 | 4 | 4 | 2 |
| DT32 | 32 | 32 | 32 | 32 | 16 | 16 | 8 | 4 | 4 | 2 |
| DT64 | 64 | 64 | 32 | 32 | 16 | 16 | 8 | 4 | 4 | 2 |

If the test of the group’s combined samples is positive, indicating that one of the individuals in the group is positive, apply a routine of repeatedly dividing the group into subgroups applying tests to the subgroups until a positive individual is identified. This is step two of the group test method.

**2**. If the first test is (+), divide the group into two subgroups of equal size. Test the combined sample from individuals in one subgroup to determine which halve contains some positive individuals. Repeat this step on the subgroup that contains the positive cases. This routine is referred to as **binary search**.

### 3.1 Binary search details

When the test result of the group’s samples is (+) we split the group into two equally sized subgroups testing one of them to decide if it contains some positive cases or if it consists entirely of negative cases. In the second situation, the second half must contain positive cases.

Continue the search on the subgroup with positive cases in the same way dividing it into two halves testing the mixed samples from one of the half-subgroups. Repeating this step we are able to find one positive case. Along the way many groups of several individual are likely to be confirmed negative with a relatively small number of tests. Any subgroup that was not tested retains its undetermined status and is returned to the general set of samples to be confirmed positive or negative.

**3**. Repeat the procedure starting with 1. on the individuals in the population for which whether they carry the disease has yet to be determined.

In this paper we consider group testing methods that follow two different interpretations of step 1 in the procedure above: methods with a fixed group size and methods where the group size changes based on the results of previous tests. Each type of method uses a slightly different definition of prevalence.

#### Fixed group size

Prevalence is defined as the probability *p* an individual in the population has COVID-19. For each level of prevalence we specify a fixed number of individuals to test their combined samples, performing binary search if it is positive. The fixed group size method we analyze in this paper is **divide and test DT**. We also consider **Dorfman’s method** as a method to compare the methods against, as well as a simpler alternative that only ever uses an individual’s samples for a maximum of two tets.

#### Dynamic group size

Prevalence is defined as the absolute number of positive cases in the test population. As testing is carried out, the confirmed positive and negative cases are set aside are both removed from the undetermined test population. Therefore, the population and the prevalence changes during the testing process. The group size depends on both the population and the prevalence. The adaptive group testing method presented in this paper is **generalized binary splitting GBS**.

In addition to studying these two classes of methods, we make modifications to the methods to take into account limitations of the devices and techniques used to test samples for COVID-19 RNA. Specifically, we study methods with limited group sizes. These are DT and GBS with maximum group *g* (DT*g* and GBS*g*). They are defined so that any time either DT or GBS specifies a group size over the capped limit *g* we take the group size to be *g*. To cover a wide range of limits of detection we consider groups of sizes 4,8,16,32 and 64.

## 4 Fixed group size method: divide and test (DT)

This method is can be thought of as a fixed group size version of the generalized binary splitting method [5].

1. If *p* > .5 then test each member of the population individually. Otherwise, let 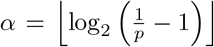. Select a group of individuals *G* with |*G*| = 2^*α*^ and apply one test to *G*. If it is negative, then we conclude that the result of each case in *G* is negative.
2. If *G* is positive we use binary search to find exactly one case that we can deduce is positive.
3. Repeat the method starting with step 1. on the yet to be confirmed portion of the population.

Note that the 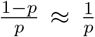. The median number of positive cases in a group 2^*α*^ is then roughly 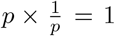, therefore the group contains only negative individuals with about the same frequency as it contains at least one positive individual.

Divide and test, although having fixed group size, can be thought of as a version of Hwang’s generalized binary splitting for large *N*.

For *N* large and *D* positive cases, the status of each case is approximately independent of the other with a probability of being positive 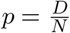. This is approximate because the generalized binary splitting algorithm assumes a fixed amount of positive cases in a fixed population and drawing cases without replacement can be approximately modeled by a sequence of independent Bernoulli random variables.

## 5 Dynamic group-size method: Hwang’s generalized binary splitting (GBS)

The group testing methods suggested in this paper are based on Hwang’s generalized binary splitting method. In [5] GBS is designed to find up to *D* (+) cases in a population containing *N* individuals. It is described as follows:

1. If 2*D* − 2 ≥ *N* then test each member of the population individually. If 2*D* − 2 < *N* set 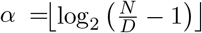. Take *G* a group of cases such that |*G*| = 2^*α*^ and apply one test to *G*. If it is negative, then we conclude that the result of each case in *G* is negative.
2. If *G* is positive we use binary search to find exactly one case that we can deduce is positive.
3. Set the population to the set of individuals not determined (+) or (−). We set *N* to be the new population size. If we have found a positive case in the last two steps, we take *D* − 1 to be the new number of positives in the un-tested population.

This method is extended to populations with a random number of (+) cases by setting the upper limit on positive cases according a chosen confidence level (i.e. the probability that the number of (−) exceeds this is very small). In probability terms, if the number of positive cases *D* is generated acccording to a probability distribution, and we assume probability *c* that will identify every positive case, then let *D*_*c*_ be such that *P* (*D* ≤ *D*_*c*_) < *c*. Generalized binary splitting is then applied to find at most *D*_*c*_ positive cases in the population.

The setting of Hwang’s paper is group testing for general purposes, including identifying defective parts or products in addition to determining individuals with a disease.

It is an ethical requirement to determine if each person is (+) or (−) if they are tested for COVID-19. Therefore GBS is appplied for an upper bound *D*_*c*_ of positive cases at a fixed level of confidence *c*. If exactly *D*_*c*_ positive cases are found, then there with non-zero probability there are some cases that have not been determined. Otherwise if < *D*_*c*_ positives are identified, then we are certain we have found them all and it will also happen that a positive or negative confirmation was given to each individual by the end of the testing process.

In the case where there could be more positive cases, we repeat GBS with the same level of confidence *c* on the remaining population.

Rather than considering the application of a level of confidence as an idiosyncracy of the method, it is actually a fundamental property of identifying asymptomatic COVID-19 carriers. Asymptomatic positives cannot be identified without testing them, we can only pick an upper bound on the cases with a high level of certainty. It might seem preferable to then choose a method that instead prescribes a group-size for each level of prevalence, such as Dorfman’s method or divide and test, but for those methods the possibility that we have misjudged the prevalence still needs to be factored in!

## 6 Confidence bounds on group testing methods

We will apply DT and GBS with groups capped at 4, 8, 16, 32 and 64 samples using two different levels of confidence.

- GBS*g* with *D*_.5_ (i.e. *P* (*D* ≤ *D*_.5_) = .5). We repeat GBS*g* at this level of confidence if there are undetermined cases remaining.
- We apply DT*g* with the group size 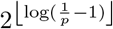.
- GBS*g* with *D*_.99_. For a population generated by a prevalence level *p*, this will perform worse on average than the application with *D*_.5_, but level of efficiency theoretically guarenteed for GBS [5] will hold with ll% confidence. Therefore, we can be more confident of the algorithm;s performance.
- For populations of size *N* generated with prevalence *p*, we apply DT*g* at the actual prevalence level of the population with 99% confidence. That is, we apply DT*g* with the group size (with ceiling *g*) corresponding to 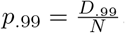.

For small to medium sized populations, the 99% confidence upper bound is very different from the mean. The different confidence levels are very close for large *N* for a constant probability of having COVID-19. How confident we are needs to be taken into account in this scenario as well because assuming a uniform probability of having the disease is likely to be too broad of an assumption. There are many different factors that lead to sub-populations having distinct frequencies of carrying COVID-19 and the certainty about the prevalence varies as well.

## 7 Dorfman’s method[3]

We compare the results from the DT and GBS methods to Dorfman’s method. Dorfman’s method is as follows:

1. For a the optimal group size *b* (defined below), apply one test to a group of *b* cases. If the result is negative, then conclude that every case in the group is negative.
2. If the result is positive, test each case individually.

If the test result is negative then one test was used for *b* cases. If it is positive, then *b* + 1 tests were used. The quantity *b* is chosen to minimize the expected number of tests used to determine the result of one individual case,

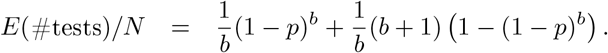

We will use this formula to compute the expected proportion of tests saved for several different levels of prevalence to compare to the successor binary search based methods we choose study.

## 8 Analysis of group testing methods methods^1^

To evaluate and analyze the performance we assume that the populations are sequences of randomly generated i.i.d. variables representing cases with the probability of having the disease *p*. The population is then modeled by a probability distribution with the following parameters.

- *N* - the size of the population/number of cases
- *p*-the probability that a case is positive. We refer to this as the prevalence.
- A case is then a Bernoulli random variable that is positive with probability *p* and negative with probability 1 − *p*.
- The number of positive cases is then a binomial random variable with *N* samples and probability *p* of a success, we write this as binomial(*N, p*).
- *T*−the number of tests needed to determine the status of every case for a population of size *N*.

Performance is assessed using two measures: the expected percentage of tests saved (compared to individual testing) and the rate information is gained from applying each test. The proportion of tests used for a sample population (compared to testing individfually) is the expression of the ratio 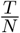. The percentage of tests saved is then 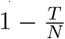 expressed as a percentage. We define the information rate following suit with the Definition 1.7 in [2] to be the number of bits gained per test. The information gained is taken to be the base 2 entropy of the sequence of the random variable of *N* independent COVID-19 (+) or (−) confirmations, denoted *H*(population). Let *H*(*p*) = *p* log_2_ *p* + (1− *p*) log_2_(1− *p*) be the entropy of a bernoulli *p* random variable. Since entropy is additive over finite collections of independent random variables, we have

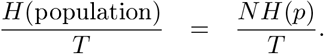

We compute the average number of bits per test by sampling populations many times and taking the mean of this quantity computed for each sample.

In section l we analyze the performance of the group testing methods at a selection of prevalences for populations of size 100 and 1000. In section 10 we analyze the performance at the .99 confidence upper bound. This can be thought of as overestimating the prevalence by a large enough amount so that we are 99% sure that the actual number of positive cases falls at or below this level.

In section 11 we analyze the number of rounds of testing each sample goes through. Our results suggest that not only do group testing methods save tests and time, but they can be done within the realistically expected amount of usage for each test.

An analysis of the histograms of the number of tests per case from which we compute the average performances is in the numerical results section after the bibliography (section 14).

## 9 Performance of group testing methods at different prevalences

For our performance analysis, we sample populations of sizes *N* = 100, 1000 at prevelance levels *p* = .001, .01, .02, .03, .04, .05, .1, .15, .2, and .25. We present the mean of 1000 samples and present them in the table below. For DT*g* results, some of the entries are left empty. This is because the fixed group size is less than the capped group size so it would be redundant to list it; for the performance refer to the nearest entry to the left. We do not do this for GBS because the group size is adaptive and there is nothing restricting it from becoming small.

First we assess the performance of DT at its uncapped group size, recording the group-size, the percentage of tests saved and the information rate.

**Table 9.1:** We sample populations with populations the same size as the group size for the DT method 10000 times. We find that the average rate of information per test [2] is approximately .96, and due to the definition of our algorithm in terms of the probability of a group testing negative being 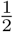, regardless of prevalence, we feel this is a good approximation for all prevalence levels. The number of bits per +/− status of a case steeply increases in the beginning and levels off at 1. This expresses the negative relationship between the effectiveness of group-testing and increasing prevalence.

| prevalence- $p$ | group-size- $2^{\lceil \log_2 \frac{1}{p} - 1 \rceil}$ | tests-saved | information rate (bits/test) | bits/case $H(p)$ |
| --- | --- | --- | --- | --- |
| .0005 | 1024 | 99.4% | .96 | .006 |
| .001 | 512 | 99% | .97 | .01 |
| .01 | 64 | 92% | .95 | .08 |
| .02 | 32 | 86% | .97 | .14 |
| .03 | 32 | 80% | .97 | .19 |
| .04 | 16 | 75% | .97 | .24 |
| .05 | 15 | 70% | .96 | .29 |
| .1 | 8 | 51.4% | .96 | .46 |
| .15 | 4 | 37% | .97 | .6 |
| .2 | 4 | 25% | .96 | .72 |
| .25 | 2 | 15.5% | .96 | .81 |

The results of 9.1 indicate the near optimality of divide and test. It recovers nearly a bit of information for each test. For low prevalence, the confirmation of each case holds a very low amount of entropy. This is a way to understand the quantitative relation between tests saved and prevalence by group testing.

In each table below we list the percentage of tests saved from employing the group test methods with capped group sizes.

**Table 9.2:** Tests saved for GBS method with capped group size. We sample populations at prevalence level *p* (left column) for sizes 100 and 1000 and simulate the GBS methods for different capped group sizes. We sample populations 1000 times each.

| prevalence | $N = 100$ | | | | | $N = 1000$ | | | | |
| --- | --- | --- | --- | --- | --- | --- | --- | --- | --- | --- |
|  | GBS4 | GBS8 | GBS16 | GBS32 | GBS64 | GBS4 | GBS8 | GBS16 | GBS32 | GBS64 |
| .001 | 74.7% | 86.7% | 92.5% | 95.4% | 96.3% | 74.7% | 87.1% | 93.2% | 96.2% | 97.6% |
| .01 | 72.2% | 83.2% | 88.0% | 90.1% | 90.1% | 72.3% | 83.6% | 88.7% | 90.6% | 91.2% |
| .02 | 69.0% | 79.2% | 82.6% | 83.5% | 83.6% | 69.8% | 80% | 84.0% | 84.8% | 84.7% |
| .03 | 66.6% | 75.1% | 77.9% | 77.9% | 77.8% | 67.3% | 76.3% | 79.2% | 79.1% | 79.1% |
| .04 | 64% | 71.2% | 73.2% | 72.6% | 72.6% | 64.8% | 72.8% | 74.5% | 74.3% | 74.2% |
| .05 | 61% | 67.5% | 68% | 67.6% | 68.1% | 62.3% | 69.0% | 69.8% | 69.8% | 69.5% |
| .1 | 47.9% | 49.3% | 49.2% | 48.7% | 48.8% | 49.9% | 51.1% | 50.8% | 50.7% | 50.6% |
| .15 | 35.5% | 34.6% | 34.3% | 34.7% | 33.7% | 37.3% | 36.9% | 36.8% | 36.7% | 36.7% |
| .2 | 23.4% | 22.2% | 21.9% | 22.2% | 21.9% | 24.3% | 24.1% | 23.9% | 24.0% | 23.9% |
| .25 | 14.7% | 13.9% | 13.6% | 13.0% | 13.7% | 16.3% | 16.2% | 16.0% | 16.0% | 16.0% |

**Table 9.3:**
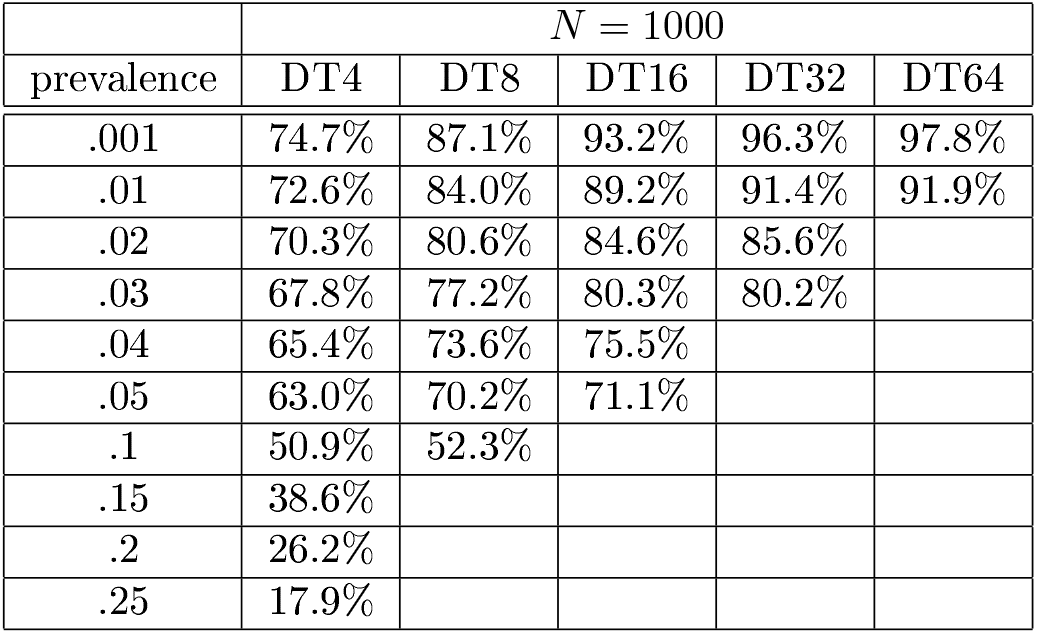
We list the performance of divide and test at different levels of prevalence. Divide and test has a constant group size given the prevalence.

Interestingly, although GBS has theoretical guarentees on the number of tests, DT has better performance on average.

**Table 9.4:** Dorfman’s method performance for different levels of prevalence. Note that, although Dorfman’s method is simpler, it requires groups more than twice the size to barely exceed the performance of GBS and DT with group size capped at 16 for prevalence .001 and capped group size at 8 for prevalence .01.

| Dorfman |  |  |
| --- | --- | --- |
| prevalence | tests saved (%) | group size |
| $p = .001$ | 93.7% | 33 |
| .01 | 80.4% | 11 |
| .02 | 72.6% | 8 |
| .03 | 66.5% | 7 |
| .04 | 61.6% | 6 |
| .05 | 56.8% | 6 |
| .1 | 40.6% | 4 |
| .15 | 27.2% | 4 |
| .2 | 17.9% | 3 |
| .25 | 8.9% | 3 |

## 10 GBS and DT at 99% confidence

We analyze the effect of picking the prescribed method for a conservatively large estimate on the prevalence. A larger estimate on the prevalence will still take advantage of the frequent occurence of many consecutive negative cases. The overestimate we use corresponds to a probability of having less positive cases of .99. We sample the populations at least 100 times and take the mean of the proportion of tests saved over all samples. Specifically for DT*g* applied to populations of 100 and 1000 we generated 100 samples. For GBS*g* applied to populations of size 100 we sampled 1000 times except for GBS16. For GBS*g* applied to populations of size 1000 we sampled 100 times except for GBS64 we sampled 1000 times. The differences in number of samples are due to the overall computational expense of simulating GBS and DT.

The mean savings of each numerical experiment are presented in the following table.

**Table 10.1:** In this table we sample the DT and GBS methods 1000 times and take the average results for number of positives less than or equal to *D*_.99_. Again, note that we leave out entries from the table for DT since the optimal group size is smaller than the maximum group size allowed.

| | $N = 100$ | | | | | | | | | |
| --- | --- | --- | --- | --- | --- | --- | --- | --- | --- | --- |
| prevalence | GBS4 | GBS8 | GBS16 | GBS32 | GBS64 | DT4 | DT8 | DT16 | DT32 | DT64 |
| .001 | 74.7% | 86.7% | 92.7% | 95.4% | 96.2% | 74.7% | 86.6% | 92.6% | 95.5% | 97.7% |
| .01 | 66.4% | 75.3% | 77.8% | 78.3% | 78.2% | 72.7% | 84.1% | 88.8% | 88.9% | 88.4% |
| .02 | 61.2% | 67.6% | 68.3% | 67.9% | 67.9% | 70.2% | 80.6% | 79.6% | 80.7% |  |
| .03 | 56.1% | 60.4% | 60.4% | 60.4% | 60.5% | 67.1% | 76.2% | 77.7% | 76.0% |  |
| .04 | 53.6% | 57.0% | 57.0% | 57.0% | 57.0% | 65.0% | 72.3% | 72.8% |  |  |
| .05 | 48.5% | 49.7% | 49.8% | 49.7% | 49.7% | 62.6% | 69.7% |  |  |  |
| .1 | 30.2% | 30.2% | 30.0% | 30.0% | 30.2% | 50.2% | 50.8% |  |  |  |
| .15 | 17.6% | 17.6% | 17.6% | 17.6% | 17.6% | 30.6% |  |  |  |  |
| .2 | 8.5% | 8.5% | 8.6% | 8.5% | 8.3% | 23.6% |  |  |  |  |
| .25 | .9% | .9% | 1.1% | 1.0% | .8% | 0.0% |  |  |  |  |

**Table 10.2:** In this table we take the average of 100 or 1000 samples of GBS and DT at a population of 1000 and various prevalences at the .99 confidence level *D*_.99_. Note that *D*_.99_ is closer to *D*_.5_for higher populations (that is 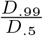 is closer to 1). Therefore the performance is closer to the results depicted in the previous section.

| | $N = 1000$ | | | | | | | | | |
| --- | --- | --- | --- | --- | --- | --- | --- | --- | --- | --- |
| prevalence | GBS4 | GBS8 | GBS16 | GBS32 | GBS64 | DT4 | DT8 | DT16 | DT32 | DT64 |
| .001 | 74.1% | 86.2% | 92.1% | 94.9% | 96.1% | 74.7% | 87.1% | 93.2% | 96.4% | 97.8% |
| .01 | 70.6% | 81.2% | 85.6% | 87.0% | 87.0% | 72.6% | 84.1% | 89.4% | 91.6% | 91.3% |
| .02 | 67.4% | 76.7% | 79.7% | 79.7% | 79.7% | 70.3% | 80.4% | 84.6% | 84.2% |  |
| .03 | 64.3% | 72.3% | 74.0% | 74.1% | 74.1% | 68.1% | 77.1% | 80.5% | 80.1% |  |
| .04 | 61.5% | 68.0% | 68.5% | 68.5% | 68.5% | 65.2% | 73.1% | 75.3% |  |  |
| .05 | 58.4% | 63.7% | 63.7% | 63.7% | 63.7% | 63.3% | 69.8% | 70.1% |  |  |
| .1 | 44.4% | 44.4% | 44.3% | 44.4% | 44.4% | 50.9% | 50.8% |  |  |  |
| .15 | 30.5% | 30.2% | 30.4% | 30.3% | 30.4% | 38.2% |  |  |  |  |
| .2 | 19.2% | 19.1% | 19.2% | 19.2% | 19.2% | 24.2% |  |  |  |  |
| .25 | 11.5% | 11.4% | 30.4% | 11.3% | 11.3% | 17.8% |  |  |  |  |

## 11 Number of uses for each sample

Although demonstrated to be more efficient in test usage, generalized binary splitting requires some samples to be used at least as many times as the logarithm of initial group size. For maximum group sizes of 32 and 64 this implies that positive cases will be tested at least 6 or 7 times. As is demonstrated by figures 11.1 and 11.2 it is possible for tests to be used more than twice as many times.

**Figure 11.1:**
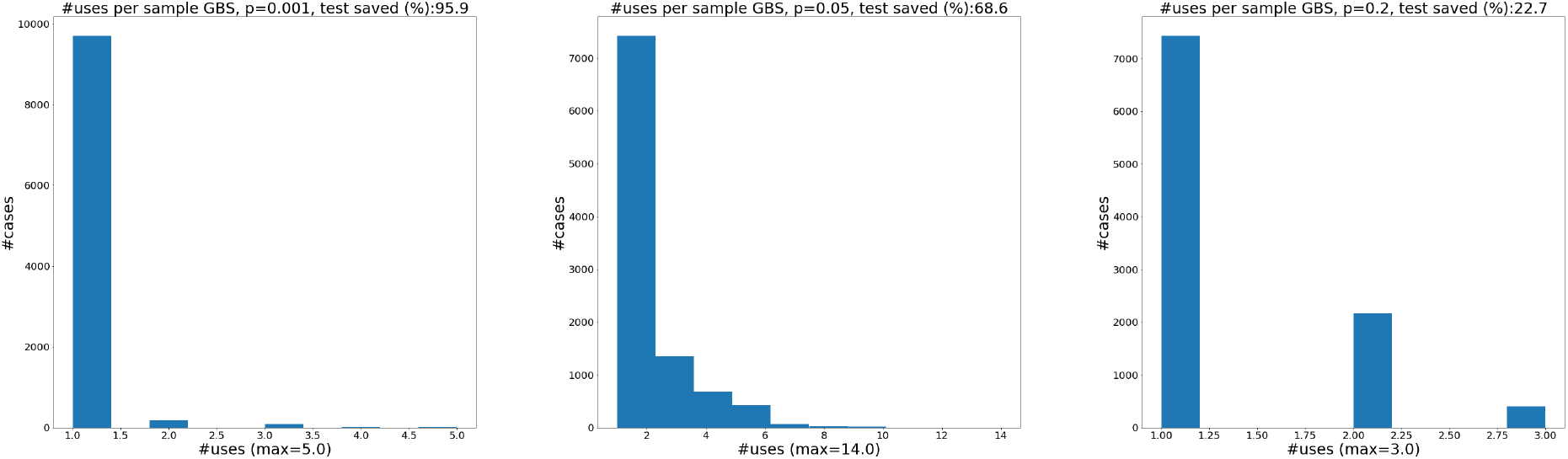
For GBS32 we sample a population of size 10,000 and record the number of times each case is tested. Note that for prevalence of *p* = .1% a sample is used at most 5 times in the population. In figure 11.2 where we perform the same experiment we find cases that are tested up to 9 times. We apply GBS32 to find at most *D*_.99_.

**Figure 11.2:**
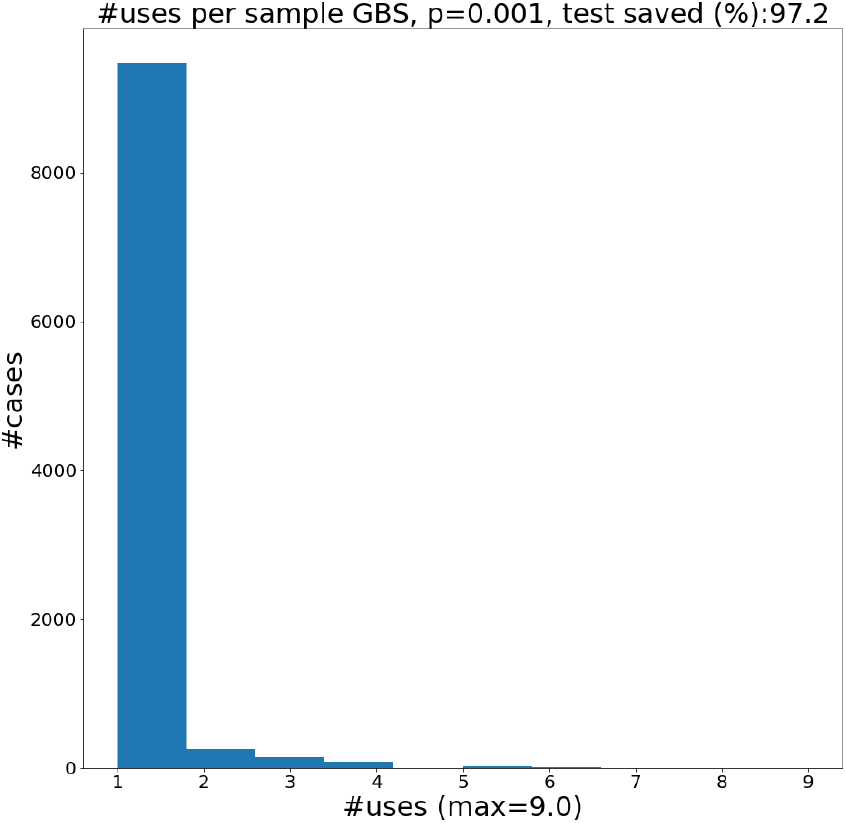
For GBS64 we sample a population of size 10000 and apply GBS64 with the .99 confidence level. The histogram shows the number of times we use each sample.

**Figure 11.3:**
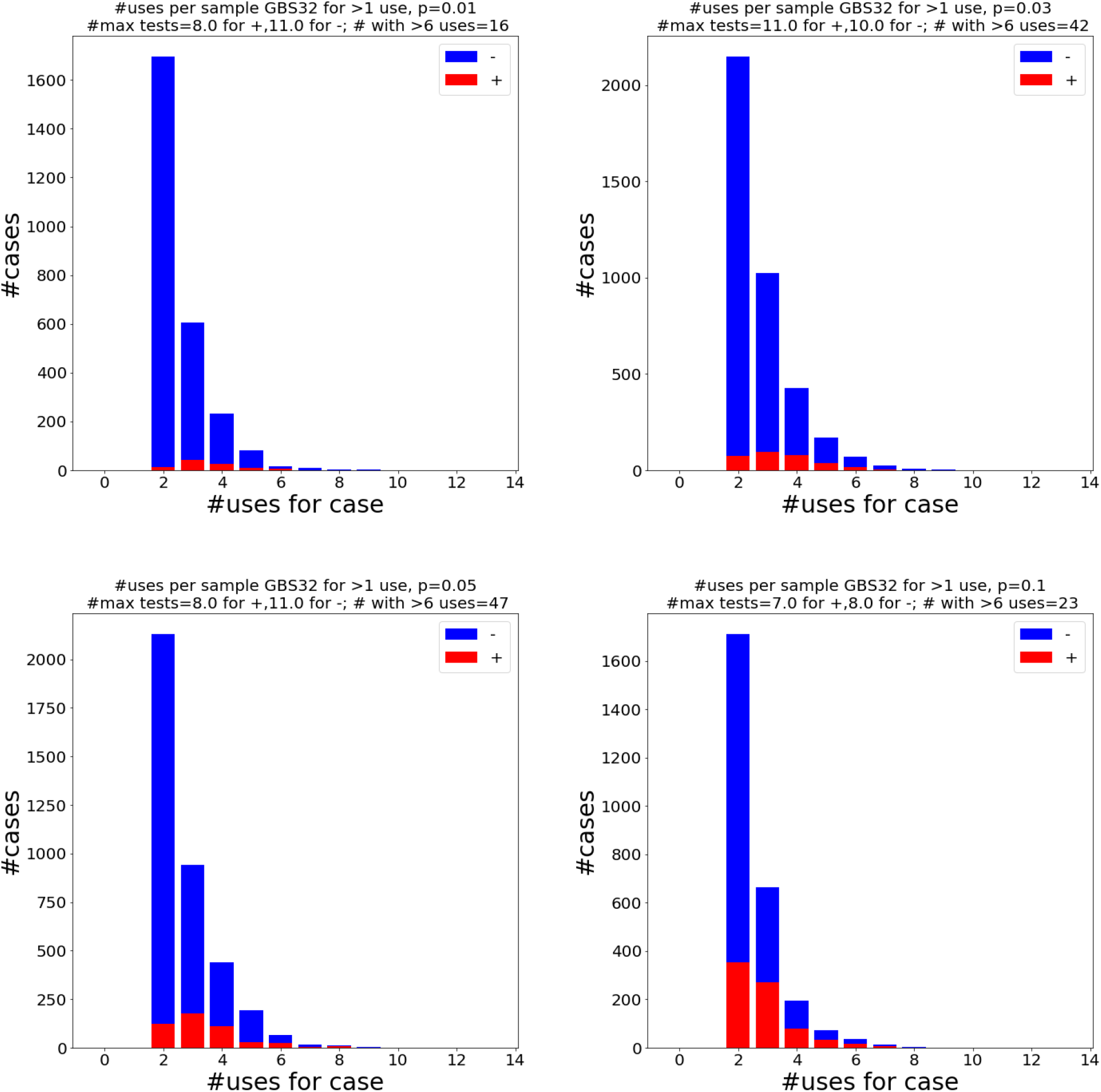
We sample a population of size 10000 at various prevalence levels and use GBS32 at the 99% confidence upper bound. We plot the histograms of the number of times postive results and negative results are tested given that they are tested more than once. Interestingly, sometimes the number of times negative cases are tested seems to have a larger maximum.

We see in figure 11.3 that the proportion of cases that are tested many times (more than 6) is very low. This is evidence that testing the cases indivually when we are running low (if we are limited to 6 uses) adds up to less than 1% extra tests. If we are able to test a sample more than this many times, individually testing when we are about to run out effects the efficiency negligibly.

## 12 Discussion

Gaining knowledge of COVID positive status in asymptomatic carriers is of prime importance in the fight to contain and eliminate the disease. The group testing strategy will generate certainty and a margin of safety in confined populations and may be useful in detection of disease burden in geographically dispersed populations as a mode of surveillance. A key factor in our strategy is the two phase approach that we propose. Clinical screening of symptomatic patients cuts down the prevalence of COVID-19 in the chosen asymptomatic test populations.

Rapid, large scale testing is urgently needed for naval ships and military bases. A recent outbreak upon the USS Roosevelt has brought this concern to the forefront of military proporities 17. Recently Italian prisoners have rioted rioting due to lack of testing1l. 300 prisoners have been released from NYC prisons due to concern for spread of coronavirus 110. Nursing homes have been sites of extensive outbreaks [11]

In the current phase of the pandemic, testing for the presence of COVID-19 targeted antibodies using viral antigens such as the ELISA assay are being used to test for both exposure and immunity. It is an important topic to explore how antibody tests can be most effectively used in tandem with PCR tests leading to the largest possible testing capacity. Antibody tests are important especially to discover whether first responders are potentially exposed and immune to the disease. As antibody tests are not yet produced enabling point of care usage, there is a scarcity of antibody testing capacity just as there is PCR testing capacity. Is it feasible to apply group testing methods to antibody assays?

Screening an asymptomatic population is challenging but important. Sensitivity of tests is paramount but also utilization of resources must be efficient. This study proposes a statistically valid mathematical model to optimize number of tests performed On chosen populations. The future direction of our work will require clinical validation with real world application of this group testing strategy.

The clinical screening of symptomatic patients out of our population is helpful but it will also make detecting virus in asymptomatic people potentially tougher because of the chosen viral load. Current RT-PCR methods are standard but have a concerning false negative rate in symptomatic patients 11. This may be further exacerbated in asymptomatic patients. Future studies may explore or consider the use of more sensitive techniques including DDPCR and addition of other sampling techniques such as fecal specimens and perhaps advanced imaging such as chest CT scan.

Based on the above evidence it is clear that a comprehensive strategy is necessary to test all asymptomatic people. This strategy will uncover the hidden silent carriers of disease.

Group testing has the chance for saving tests, while giving an exit to revert to individual testing if there are more cases than estimated. These strategies are not necessarily the theoretical optimums. Nevertheless, by our calculation and numerical simulations they have the power to cut down the number of tests used.

## 13 Conclusion

The group testing method is applicable to confined populations from both a clinical and mathematical stand point. We can clinically screen confined groups to decrease the prevalence in a predictable way. This method allows each patient in the population to get a test result. We explore adaptive and non-adaptive strategies for group testing for COVID-19, therefore we add adaptive and binary search based non-adaptive strategies. Both perform well even when we restrict the size of the group based on the sensitivity of RT-PCR.

We test the performance of two algorithms: divide and test and generalized binary splitting. For populations of size 100 and 1000 with each case being positive independent of one another with probability *p*, we find that both methods save many tests. In fact, even when we restrict the size of the groups they make substantial savings. For example at prevalence .001 and group sizes capped at 16, both DT16 and GBS16 are capable of outperforming Dorfman’s method, that requires a group size of 33 for optimal performance. For a population of 1000 and confidence level .99, the DT16 and GBS16 also have a similar performance to Dorfman’s method at .001 where Dorfman’s method is chosen as if we knew the exact prevalence.

We find that DT outperforms GBS on average even though GBS in theory uses close to optimal number of tests for a known fixed number of (+) cases (bounded above by #tests − 1 + information lower bound). This indicates that methods based on DT have the potential to save many tests. We found that DT when applied without a capped recovers close to the optimal 1 bit of information with each group-test. If the measurement is sensitive to detect one positive in groups of size twice up to 16 as large as the current maximum, then optimally efficient testing can be carried out. It might be the case that a full binary search of the group puts too much strain on the laboratory work flow. The bit of information that gained from negative test results on large pools can be leveraged while choosing smaller sample sub-groups to carry out the process of finding positive cases.

Since samples can be divided, frozen and stored for continuous use we mathematically analyze how many divisions we have to use for each sample. We find that testing individually when the sample is about to run out does not subtract substantially from the savings. We demonstrate that for a group size of 32 the generalized binary search on 10000 cases uses the same sample more than 6 times for < 1% of all cases. Therefore, applying a test individually to samples when they are running low is a viable way to save many tests while not depleting material from one person’s sample.

## Data Availability

Jupyter Notebook for numerical experiments may be found at https://github.com/cmentus/group-testing/.

https://github.com/cmentus/group-testing/

## Acknowledgements

We thank 1artin Douglas for interesting and useful discussions about group testing methods using binary search. We are also grateful to Matthew Heidemann, Blake Elias, Gary Chizever, 1ichael Wells, Olga Buchel and Patrick O’Neill for contributing ideas and useful feedback. We thank 1atthew Heidemann for the diagram at the beginning of the paper that skillfully explains our process. We are very thankful to Yaneer Bar-Yam for important recommendations that greatly improved the ideas, clarity and organization of the paper.

We are grateful to Irving Epstein and Kevin Schallert for highlighting important practical considerations about false-positive and false-negative rates of PCR tests and contributing ideas about applying group testing in tandem with other methods (e.g. antibody testing). Thank you to David F. Schaeffer, 1arcel Dvorak, Julia Naso and Marthe Kenny Charles for discussions about the limit of detection for SARS-CoV-2 RNA by RT-PCR and recommendations that led us to simulate group tests with maximum sizes 16, 8 and 4. We are very thankful to Oliver Johnson for discussing information theoretic aspects of our problem and recommending the information rate as a measure of performance.

We thank EndCoronavirus.org and the New England Complexity Institute for giving us the platform to meet and collaborate as a team and meet great people with diverse backgrounds and skillsets united in the cause to fight the current pandemic.

## 14 Numerical results

### 14.1 GBS and DT at different levels of prevalence (.5 confidence)

**Figure 14.1:**
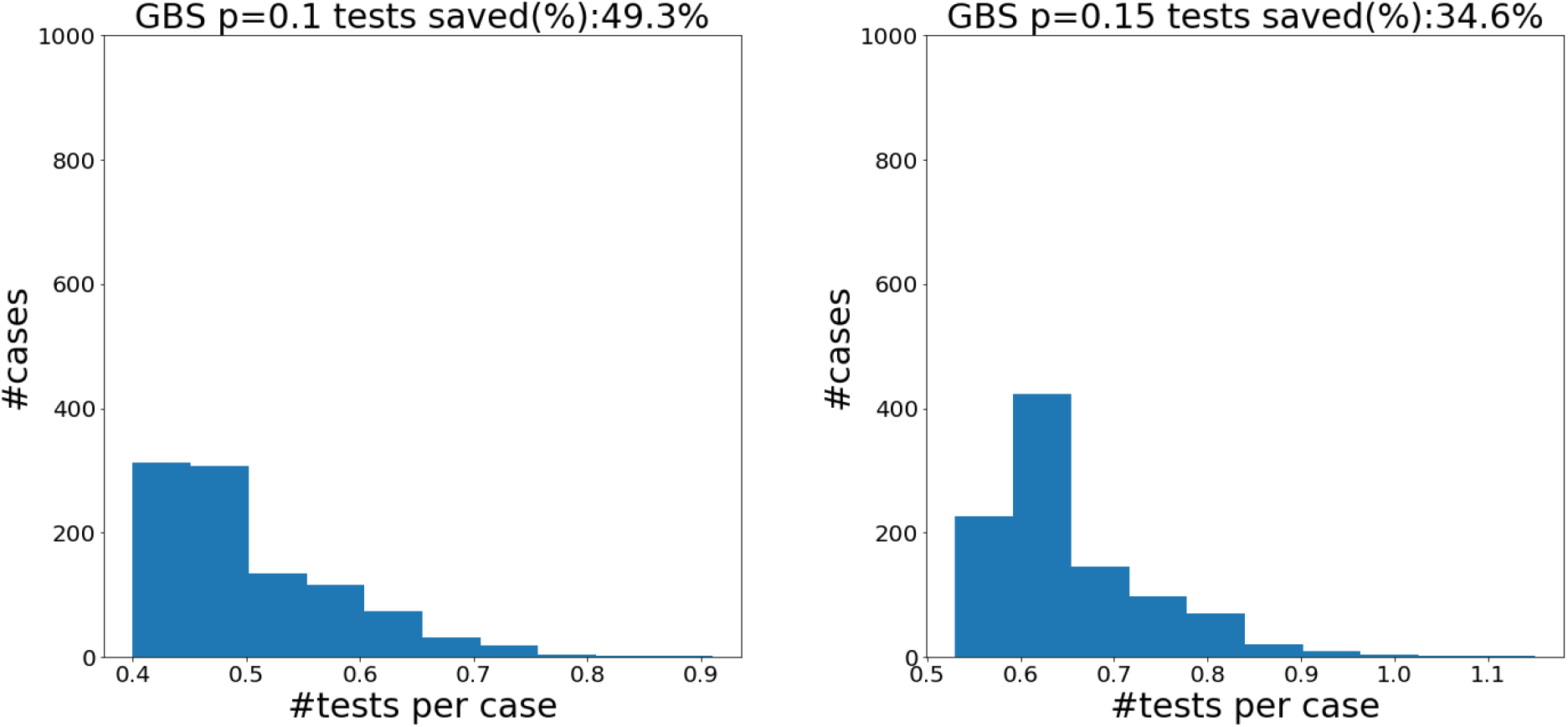
Histograms for GBS with group size capped at 8 for *N* = 100 at prevalence levels *p* = .1 and *p* = .5 sampled 1000 times.

**Figure 14.2:**
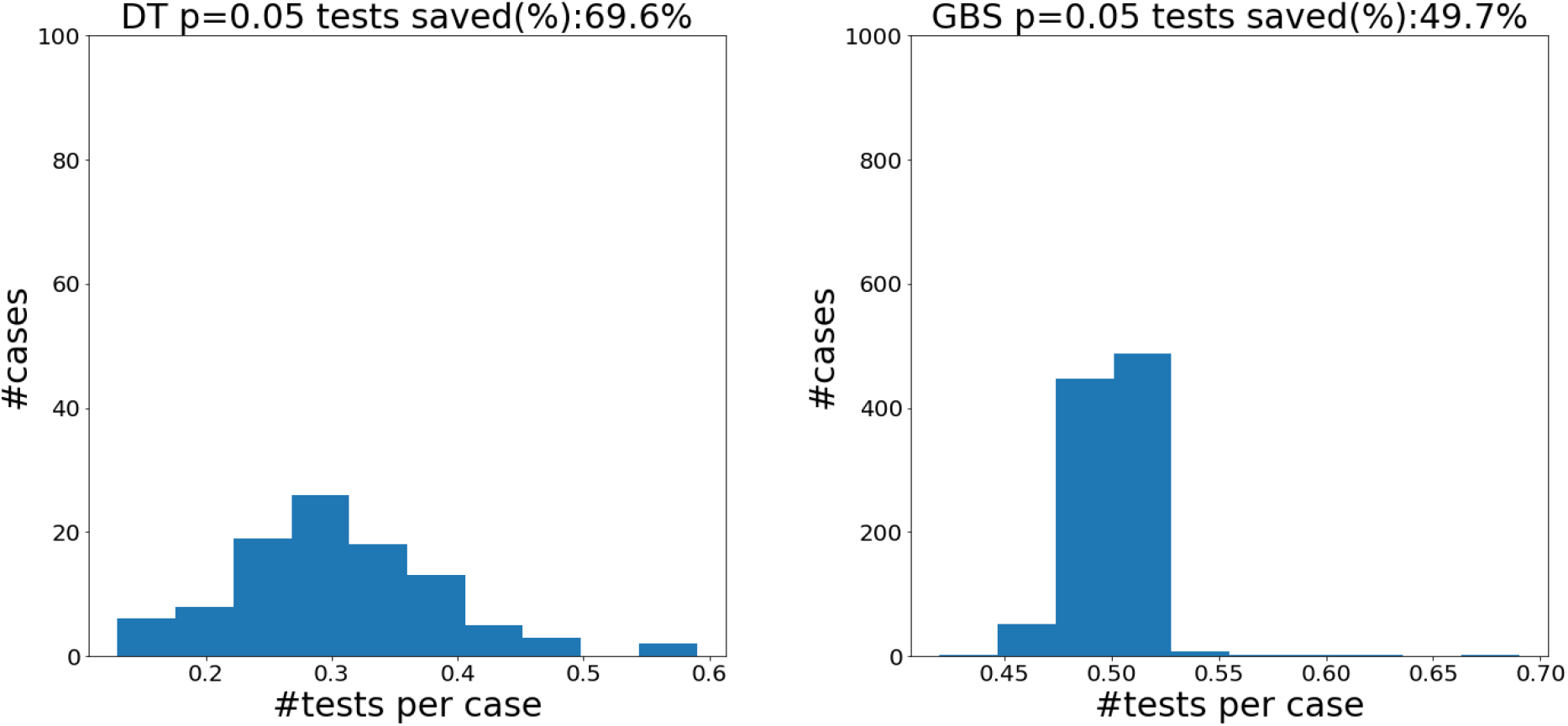
Distribution of tests per case for DT32 and GBS32 on populations of size 100 with prevalence of .5. The distribution of GBS32 is much more concentrated at .5 tests per case.

**Figure 14.3:**
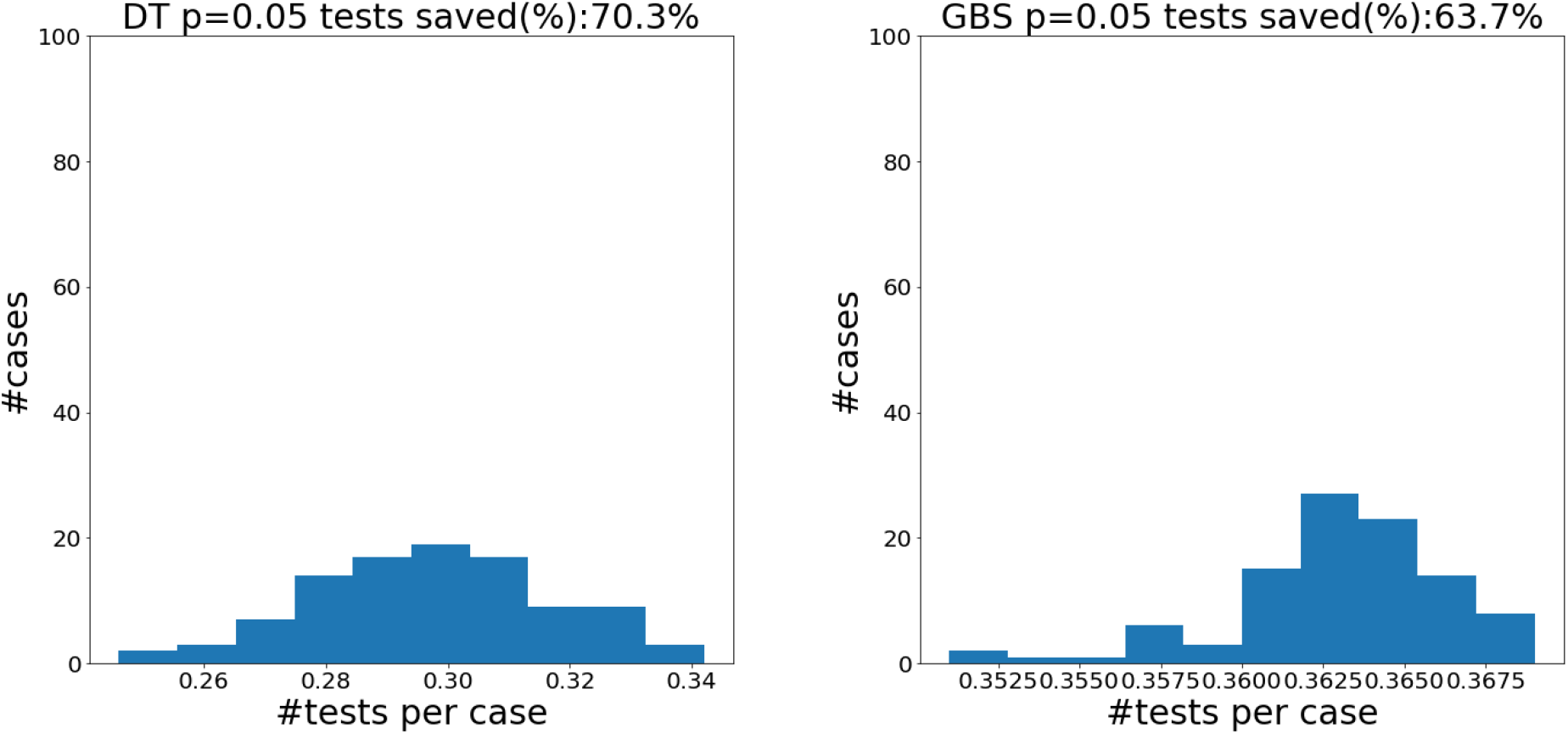
Distribution of tests per case for DT32 and GBS32 on populations of size 1000 with prevalence .05. Again, the performance of GBS is tightly concentrated at its mean.

**Figure 14.4:**
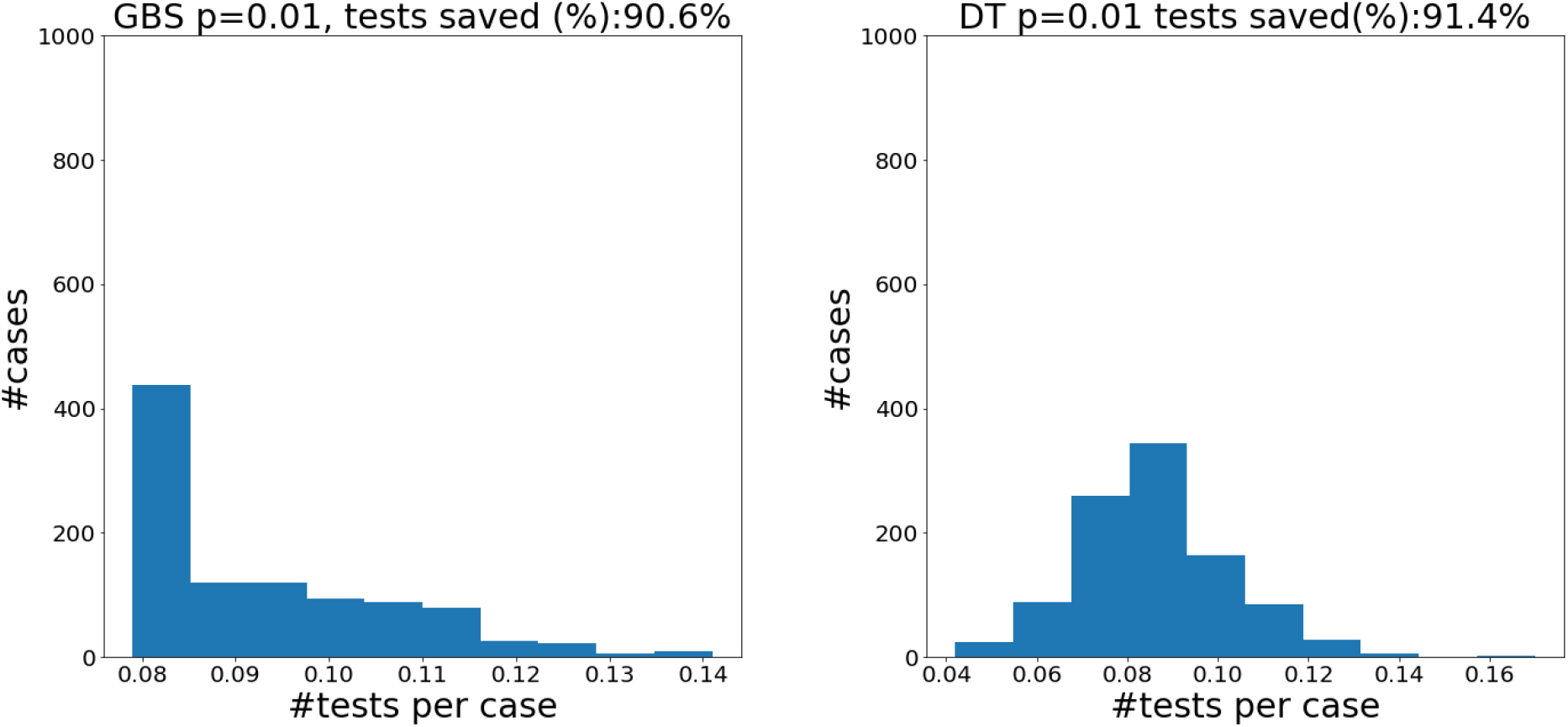
Tests per case. Left : DT32 on a population of size 1000. Right: GBS32 on a population of 1000. The prevalence of both populations is *p* = .01. The results are for DT32 and GBS32 performed assuming the .5 confidence upper bound.

### 14.2 GBS and DT at different levels of prevalence with .99 confidence

**Figure 14.5:**
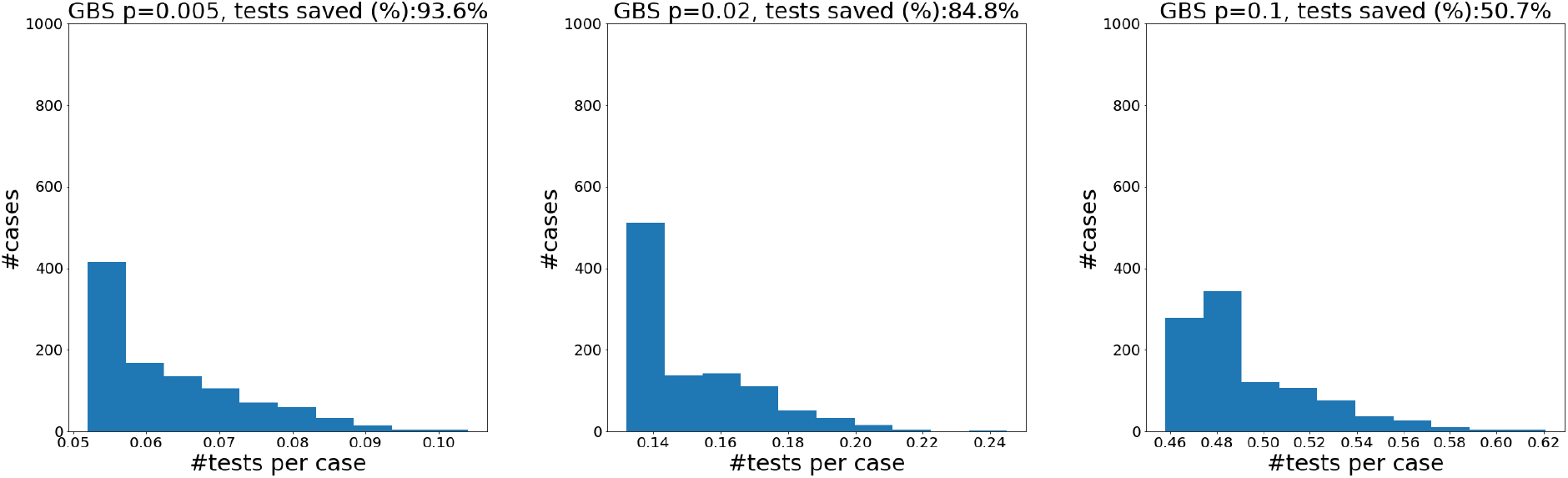
Histograms of the number of tests per case for GBS32 with population of size 1000 sampled 1000 times.

**Figure 14.6:**
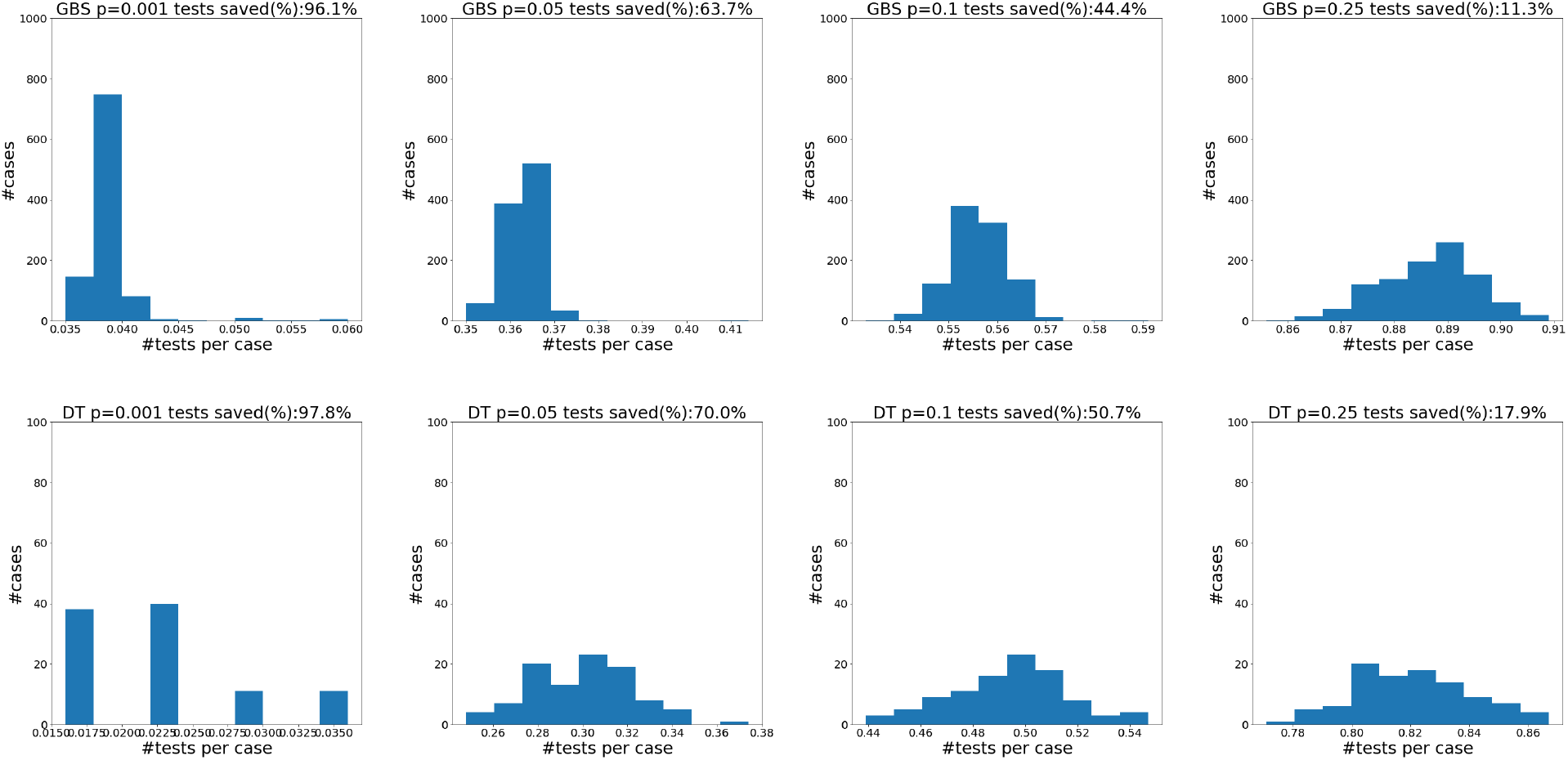
Histograms of the number of tests per case for populations of size 1000 at confidence level .99 for both GBS and DT. The maximum group size is 64. For GBS we sampled 1000 populations, for DT we sampled 100 populations. Note that the spread of the DT algorithm is much wider and has many cases that need a much lower number of tests than GBS. The small bars in GBS are caused by the 1% chance of not identifying all of the cases in one run and so repeating the process on the rest of the population.

## Footnotes

1 Please visit https://github.corn/crnentus/group-testing/ to find the Jupyter Notebook for all numerical experiments.

